# Projected health and economic impact of PCV20 vaccination in UK children: a dynamic transmission model

**DOI:** 10.64898/2026.05.12.26352641

**Authors:** Sophie Warren, Jack Said, Joe Trim, Euan Dawson, Michele Wilson, Benjamin M Althouse, Mark Rozenbaum

## Abstract

**Background:** Despite the significant impact of longstanding paediatric pneumococcal conjugate vaccine (PCV) use in the United Kingdom (UK), pneumococcal disease burden remains substantial and is primarily driven by nonl1lPCV13 serotypes. Higherl1lvalent vaccines such as the 20l1lvalent PCV (PCV20) may provide additional public health and economic benefits, yet their value in the contemporary UK setting has not been fully assessed using recent data.

**Methods:** We updated an agel1lstructured dynamic transmission model using postl1lCOVIDl1l19 UK epidemiology (2001–2023) to compare pediatric PCV20 with PCV13 and PCV15. Over a 10-year horizon, we assessed cost-effectiveness and number needed to vaccinate (NNV), capturing invasive and non-invasive disease cases, deaths, costs, quality-adjusted life-years, and incremental cost-effectiveness ratios. PCV20 was evaluated under 1+1 and 2+1 schedules; PCV13 and PCV15 were assessed under 1+1. Scenario analyses examined key uncertainties.

**Results:** PCV20 was estimated to avert more cases and deaths than PCV13 or PCV15, driven by broader serotype coverage and indirect effects. Both PCV20 schedules were dominant or cost-saving versus lower-valent comparators, with lower NNVs. PCV20’s higher vaccination costs were offset by reductions in downstream healthcare expenditures.

**Conclusion:** Paediatric PCV20 implementation in the UK could deliver substantial health gains while improving economic efficiency, supporting timely adoption.

## Introduction

In the United Kingdom (UK), pneumococcal conjugate vaccines (PCVs) have been included in the National Immunisation Programme (NIP) since 2006, originally with the seven valent PCV (PCV7) followed by the 13-valent PCV-(PCV13) in 2010 [1]. These vaccines have substantially reduced invasive pneumococcal disease (IPD) through both direct protection and indirect effects mediated by reduced nasopharyngeal carriage and transmission [2]. However, pneumococcal disease remains a burdensome infection in the UK, driven largely by serotypes not included in PCV13. Recent surveillance data reported that non-PCV13 IPD incidence was more than double that of PCV13 IPD incidence in children <1 year of age (4.4 per 100,000 vs 11.48 per 100,000 in 2022/2023), with non-PCV13 serotypes such as 8, 22F, and 9N accounting for 17%, 8% and 6% of all IPD cases, respectively [3].

In recent years, the UK paediatric PCV schedule has undergone important changes. In 2020, the Joint Committee on Vaccination and Immunisation (JCVI) transitioned PCV13 from a 2+1 schedule (2 doses before 12 months + a booster at 12 months) to a reduced 1+1 schedule, with further adjustments to the timing of the priming dose implemented in 2025[1]. Early evidence from the 2020 schedule change suggests that the 1+1 schedule may maintain population-level protection against IPD, though there has been a recent increase in both PCV13 vaccine-type and non-vaccine type disease [3]. Pneumococcal vaccines with expanded valency, including the 20-valent PCV (PCV20) and 15-valent PCV (PCV15), now have marketing authorisation in the UK and have the potential to further reduce this disease burden across all age groups.

While PCV20 has the broadest serotype coverage among licensed paediatric PCVs, the JCVI only recommends PCV13 and PCV15 for paediatric vaccination. Previous modeling evidence from Choi et al. [4] suggests that switching from PCV13 to PCV20 in the UK would significantly reduce IPD cases, while replacing PCV13 with PCV15 in children could result in an overall increase in cases. Another study which estimated the health and economic impact of switching the UK paediatric NIP to PCV20 demonstrated that PCV20 was cost-saving compared to both PCV13 and PCV15, reducing disease while saving more costs[5]. However, both modelling studies relied on pre-COVID data for model calibration, which does not reflect the current pneumococcal epidemiology in the UK. Additionally, neither of these assessments accounted for the recent change to the timing of the priming dose from 3 to 4 months, which may have implications for protection in the first year of life. Accordingly, updated assessments of the health and economic effects of PCV20 and PCV15 are needed to reflect the dosing schedule change and available data.

In addition to cost-effectiveness arguments, factors such as the number needed to vaccinate (NNV) are also becoming important considerations for comprehensive decision-making. NNV offers a transparent and intuitive measure of population-level impact and efficiency. As vaccine policy decisions in the UK are often made with consideration to finite healthcare budgets and competing social priorities, assessing alternative value estimates for pneumococcal vaccines beyond cost-effectiveness can aid vaccine prioritisation. Recently, the JCVI leveraged NNV calculations to inform considerations related to the benefits of continued COVID-19 vaccination, resulting in a move towards booster programs that were more targeted to specific populations rather than universally administered [6,7]. In the context of pneumococcal vaccines, this additional estimate of value can complement cost-effectiveness for decision-making purposes.

Therefore, this analysis aims to deliver a comprehensive assessment of PCV20’s potential value. Specifically, this study will (1) assess the cost-effectiveness of PCV20 in a 2+1 and 1+1 dosing schedule relative to PCV13 and PCV15 in a 1+1 dosing schedule, using the most recent epidemiological and cost data in the UK to ensure relevance and accuracy, and (2) determine the number of children that need to be vaccinated with PCV13, PCV15, and PCV20 to prevent a single case of pneumococcal disease or avoid a pneumococcal-related death, offering a clear metric for public health impact.

## Methods

### Dynamic model structure

We re-calibrated a previously published dynamic transmission model from the UK to the most recently available pneumococcal epidemiology data [5]. The model estimated pneumococcal disease incidence following PCV20, PCV15, and PCV13 implementation and the associated economic burden of each disease state. The epidemiologic component is a deterministic, age-structured transmission model that projects pneumococcal carriage, disease progression, and recovery within the population over a specified time horizon. The model has been previously described [5] and the model structure is presented in Figure 1.

**Figure 1.**
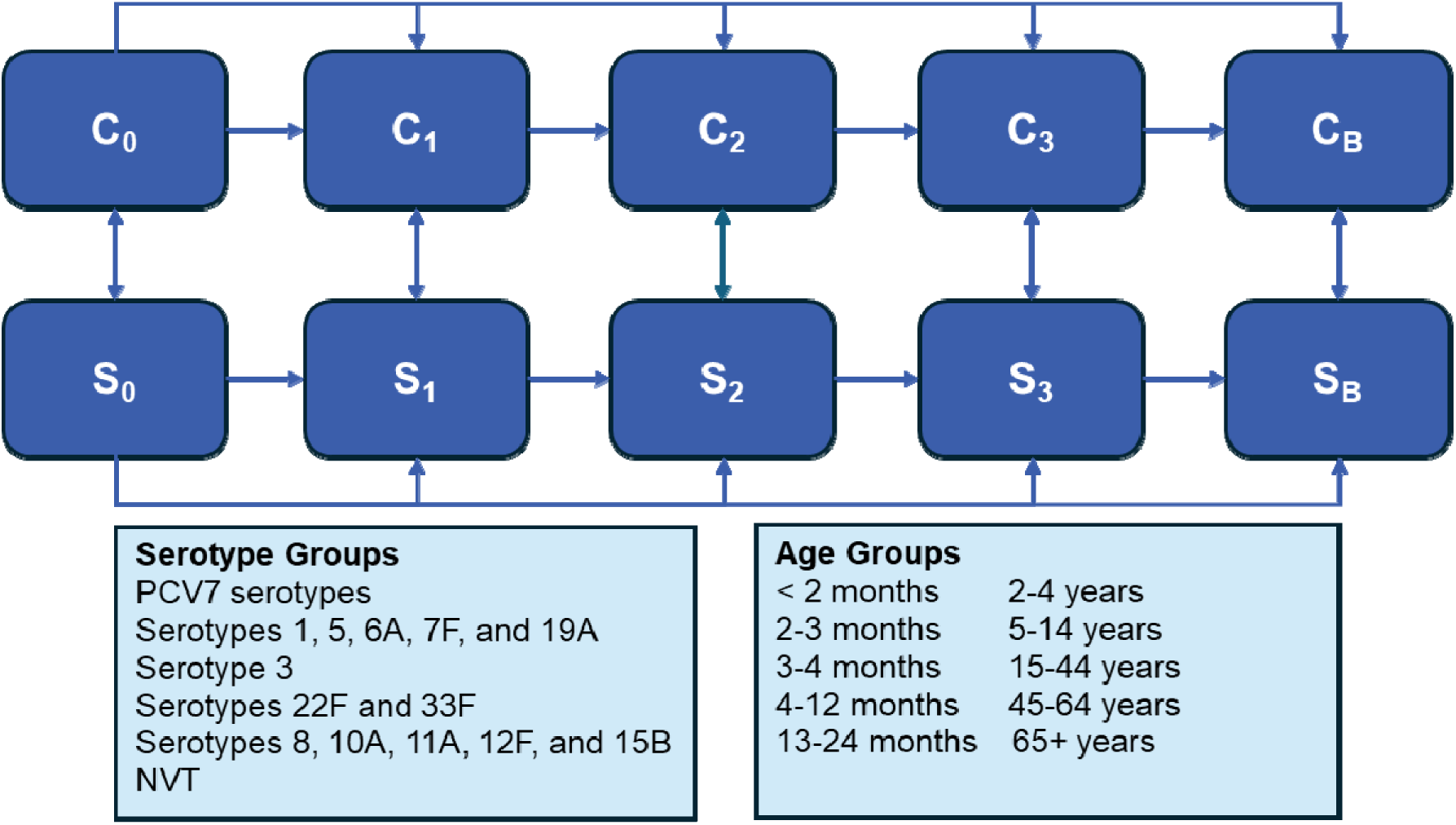
Model structure. C = carrier; NVT = non-vaccine type; S = susceptible. Note: Subscripted numbers indicate the vaccine primary dose number received; the subscript “B” indicates the booster dose received. Compartments of carriage are stratified based on development of IPD or not. Compartments relevant to vaccine doses (both carriage and susceptible) are further stratified into “no immunity” and “full immunity” which is determined by vaccine effectiveness, age group, and serotype group.

The model leveraged a susceptible–infected–susceptible (SIS) structure. Individuals entered the model as susceptible and could become carriers of a single pneumococcal serotype following contact with another infectious carrier. The risk of acquiring carriage is dependent on the serotype carried, the degree of immunity offered from prior vaccination, and the number of frequency of contacts with carriers. Upon acquisition of a serotype, a proportion of carriers developed IPD, after which they were treated and re entered the susceptible compartment, or they were subject to disease-related mortality and removed from the model. Therefore, pneumococcal transmission was dictated by carriage prevalence rather than clinical disease incidence alone. To limit model complexity, the model allowed carriage of only one serotype / serotype group at a time. Additionally, no cross-serotype immunity was assumed. Over time, individuals cleared the serotype and returned to the susceptible state. Immunity from vaccination would thus increase the opportunity for serotype replacement, as fewer cases of carriage of vaccine-covered serotypes would allow for carriage of non-covered serotypes to increase.

The model tracked vaccination history using discrete compartments corresponding to no vaccination, partial primary series (where applicable), completed primary series (where applicable), and receipt of the booster dose. Vaccination status is stratified by age, vaccine, serotype groups protected against (dependent on which vaccine an individual received), and immunity status (dependent on the number of doses received). Individuals transitioned between compartments based on vaccination schedule, uptake rates, waning, age, and infection status.

In addition to projections of pneumococcal carriage and IPD, the model estimated noninvasive disease (hospitalised pneumonia and pediatric otitis media [OM]) incidence. Specifically, hospitalised pneumonia and OM incidence was assumed to vary as a proportional ratio of IPD incidence. The model conservatively excluded outpatient pneumonia. The clinical outcomes of the model included disease cases, disease-related deaths, and life-years across the entire population.

#### Calibrated parameters

The model calibrated age– and serotype-specific IPD incidence to historical surveillance data from England and Wales collected from 2001 to 2023. Briefly, the model calculated effectiveness against IPD as a function of effectiveness against overall IPD (VE_OI_) and the effectiveness against carriage (VE_c_), where *VE_IPD_* is equivalent to vaccine effectiveness against IPD given carriage:

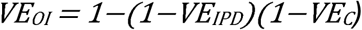

Vaccine effectiveness against PCV13 serotypes was assumed to be the same across vaccines. Due to a lack of data regarding effectiveness against IPD or carriage for PCV15 and PCV20, the model assumes newly covered serotypes in these vaccines have the same effectiveness as PCV13 did against serotypes 1, 5, 7F, 6A, and 19A. Vaccine effectiveness for PCV13 was calibrated to historical IPD data and then visually inspected to ensure calibrated estimates were comparable to published literature [8].

Calibrated vaccine effectiveness estimates can be found in Tables S3 and S4 of the supplemental material.

Additional parameters calculated during the calibration process included the probability of serotype carriage acquisition following exposure to a carrier in the absence of vaccination, the average duration of carriage, the probability of developing IPD given carriage acquisition, as well as the mean duration of protection provided by each vaccine dose.

#### Economic outcomes

The economic component applied direct medical costs and health utility decrements to modeled disease outcomes. Outcomes included vaccination costs, disease-related costs, quality-adjusted life-years (QALY), as well as the incremental cost-effectiveness ratio (ICER) when comparing each strategy to another.

#### Time horizon and discounting

Under each strategy, the model estimated IPD incidence (defined as bacteremia and meningitis) for the entire UK population over a 10-year time horizon. Costs and outcomes were discounted annually at 3.5% per year.

### Model inputs

Population, epidemiology, cost, and utility inputs are summarised in Table 1.

**Table 1.**
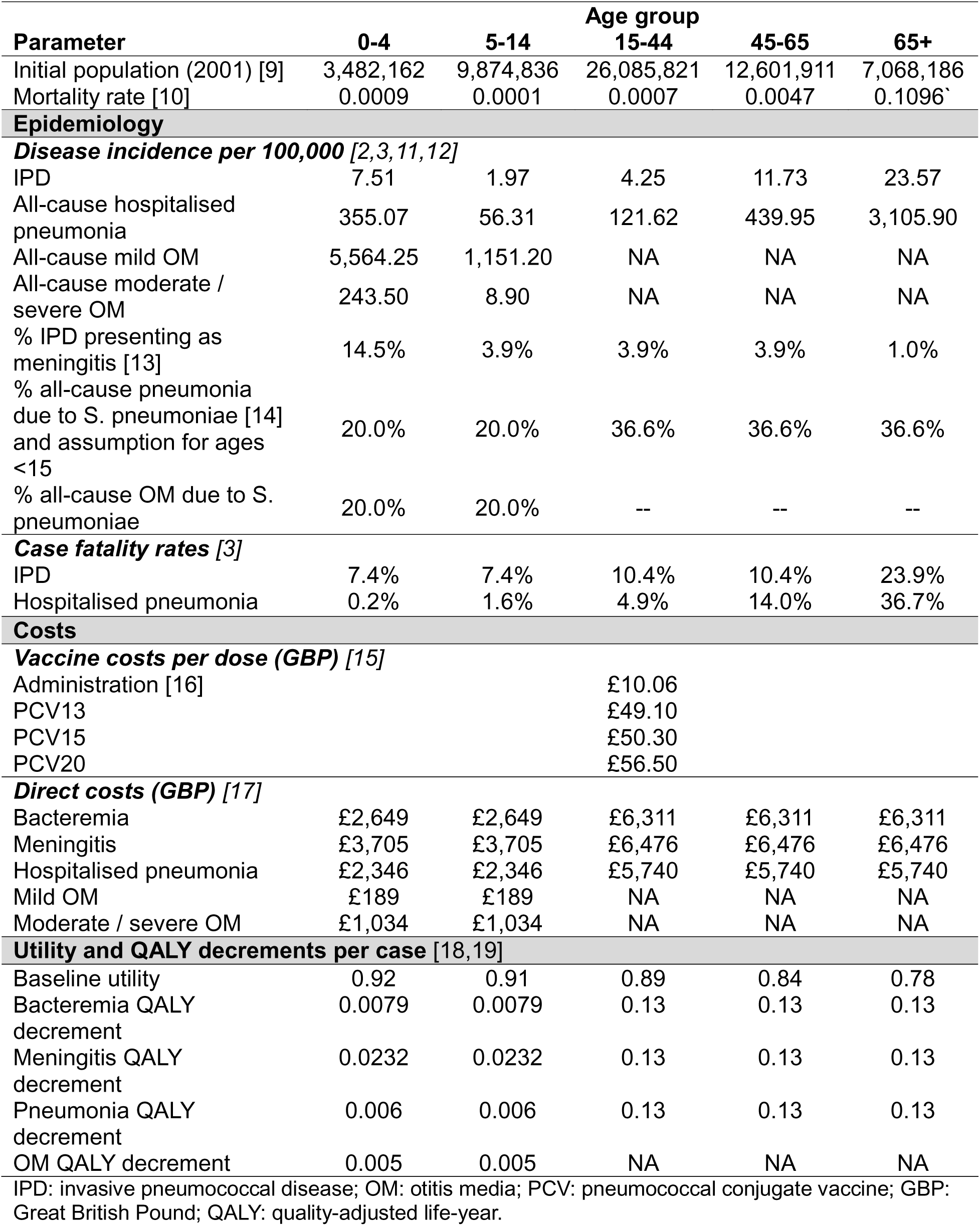
Model inputs.

| Parameter | Age group |  |  |  |  |
| --- | --- | --- | --- | --- | --- |
|  | 0-4 | 5-14 | 15-44 | 45-65 | 65+ |
| Initial population (2001) [9] | 3,482,162 | 9,874,836 | 26,085,821 | 12,601,911 | 7,068,186 |
| Mortality rate [10] | 0.0009 | 0.0001 | 0.0007 | 0.0047 | 0.1096` |
| **Epidemiology** |  |  |  |  |  |
| ***Disease incidence per 100,000 [2,3,11,12]*** |  |  |  |  |  |
| IPD | 7.51 | 1.97 | 4.25 | 11.73 | 23.57 |
| All-cause hospitalised pneumonia | 355.07 | 56.31 | 121.62 | 439.95 | 3,105.90 |
| All-cause mild OM | 5,564.25 | 1,151.20 | NA | NA | NA |
| All-cause moderate / severe OM | 243.50 | 8.90 | NA | NA | NA |
| % IPD presenting as meningitis [13] | 14.5% | 3.9% | 3.9% | 3.9% | 1.0% |
| % all-cause pneumonia due to *S. pneumoniae* [14] and assumption for ages <15 | 20.0% | 20.0% | 36.6% | 36.6% | 36.6% |
| % all-cause OM due to *S. pneumoniae* | 20.0% | 20.0% | -- | -- | -- |
| ***Case fatality rates [3]*** |  |  |  |  |  |
| IPD | 7.4% | 7.4% | 10.4% | 10.4% | 23.9% |
| Hospitalised pneumonia | 0.2% | 1.6% | 4.9% | 14.0% | 36.7% |
| **Costs** |  |  |  |  |  |
| ***Vaccine costs per dose (GBP) [15]*** |  |  |  |  |  |
| Administration [16] |  |  | £10.06 |  |  |
| PCV13 |  |  | £49.10 |  |  |
| PCV15 |  |  | £50.30 |  |  |
| PCV20 |  |  | £56.50 |  |  |
| ***Direct costs (GBP) [17]*** |  |  |  |  |  |
| Bacteremia | £2,649 | £2,649 | £6,311 | £6,311 | £6,311 |
| Meningitis | £3,705 | £3,705 | £6,476 | £6,476 | £6,476 |
| Hospitalised pneumonia | £2,346 | £2,346 | £5,740 | £5,740 | £5,740 |
| Mild OM | £189 | £189 | NA | NA | NA |
| Moderate / severe OM | £1,034 | £1,034 | NA | NA | NA |
| ***Utility and QALY decrements per case [18,19]*** |  |  |  |  |  |
| Baseline utility | 0.92 | 0.91 | 0.89 | 0.84 | 0.78 |
| Bacteremia QALY decrement | 0.0079 | 0.0079 | 0.13 | 0.13 | 0.13 |
| Meningitis QALY decrement | 0.0232 | 0.0232 | 0.13 | 0.13 | 0.13 |
| Pneumonia QALY decrement | 0.006 | 0.006 | 0.13 | 0.13 | 0.13 |
| OM QALY decrement | 0.005 | 0.005 | NA | NA | NA |

IPD: invasive pneumococcal disease; OM: otitis media; PCV: pneumococcal conjugate vaccine; GBP: Great British Pound; QALY: quality-adjusted life-year.

#### Population and uptake

Age-specific population from 2000/2001 was obtained from the [9]. The model stratified the age groups by <5, 5–17 years, 18–49 years, 50–64 years, and ≥65 years, though more granular age groups were presented in the results to capture differences in dosing schedules.

Vaccine uptake for all vaccines was based on National Health Service (NHS) data and assumed to be the same as reported uptake for PCV13 (93% uptake for priming doses and 88% uptake for booster doses)[20]. The model assumed only infants <2 years old were eligible for vaccination. The impact of an adult programme in the model was anticipated to be minimal and was therefore not included in the analysis, consistent with the previous approach[5]. In a 1+1 schedule, infants received the priming dose at 4 months and the booster dose at 12 months, whereas in a 2+1 schedule infants received the priming doses at 2 and 4 months and the booster dose at 12 months. General mortality rates for the population were obtained from national vital statistics [10].

#### Epidemiology inputs

##### Disease incidence and presentation

Historical IPD incidence from 2001-2018 was obtained from Ladhani et al[2], with more recent data from 2019-2023 obtained from Bertran et al[3]. The model assumed IPD presented as meningitis in 14.5%, 3.9%, and 1.0% of cases in individuals aged <5, 5-64, and 65+, respectively, based on UK meningitis surveillance data from 2016 [13].

Due to limited data on the incidence of hospitalised pneumococcal pneumonia and OM in the UK, the model used all-cause incidence rates and assumed a proportion of all-cause disease was pneumococcal. The incidence of all-cause, hospitalised, community-acquired pneumonia for all ages was obtained from UK Hospital Episode Statistics (HES) data [12]. Mild and moderate/severe OM incidence was obtained from The Health Improvement Network (THIN) database for children aged <18 years old [11]. The model assumed 20.0% of pneumonia and OM cases in children were pneumococcal as an assumption in the absence of published data, as well as 36.6% of adult pneumonia cases were pneumococcal, based on published literature [14].

##### Case-fatality

Case fatality rates (CFRs) for IPD and pneumococcal pneumonia were estimated using published literature. For IPD, CFRs ranged from 7.4% in children to 23.9% in adults older than 65 [4]. Pneumonia CFRs ranged from 0.2% in children less than 5 to 36.7% in adults older than 65.

##### Contact matrices

Contact matrices were obtained from Choi et al [4], which leveraged a combined contact matrix that used the number of physical contacts between participants and contact persons from the GB POLYMOD [21] and an infant survey from van Hoek et al[22].

#### Economic inputs

##### Direct medical costs per manifestation

Average unit costs for bacteremia, meningitis, hospitalized pneumonia, and OM were obtained from the 2022/2023 National Schedule of NHS Costs [17] to align with the date of the latest model-calibrated IPD incidence data from Bertran et al. [3].

International Classification of Diseases, Tenth Revision (ICD-10) codes for each manifestation were mapped to relevant Healthcare Resource Group (HRG) codes and, for each HRG code, activity-weighted national average unit costs per finished consultant episode (FCE) were derived. Where multiple HRG codes were relevant for a single manifestation, a weighted average of the individual HRG costs was taken to generate a single value. Unit costs for bacteremia, meningitis, and pneumonia were reported separately for adults and pediatrics, based on their unique HRG codes. The costs for OM were reported for pediatrics only and broken down into moderate/severe cases (admitted attendance) and mild cases (outpatient attendance).

For each manifestation, the average cost per hospital admission was then calculated by multiplying the average unit cost per FCE by the average number of FCEs per admission, derived from NHS Digital’s Hospital Admitted Patient Care Activity 2022-23 [23]. Further details are provided in the supplementary material.

##### Vaccine costs

Vaccine prices were obtained from the British National Formulary, with the price of PCV13, PCV15, and PCV20 set at £49.10, £50.30, and £56.50, respectively [15]. The cost of vaccine administration was £10.06, based on administration fees obtained from the NHS [16].

#### Utility inputs

Age-specific baseline utility estimates for the UK were sourced from Ara and Brazier[18] and QALY decrements for each disease state were obtained from Rozenbaum et al[19].

### Model analyses

Two analyses comparing PCV20, PCV13 and PCV15 were conducted, including a 10-year cost-effectiveness analysis and an estimation of the number needed to vaccinate (NNV).

#### Cost-effectiveness analysis

The base-case cost-effectiveness analysis was conducted from a UK payer perspective over a 10-year time horizon. Cost-effectiveness was assessed at willingness-to-pay thresholds of £20,000, £25,000, and £30,000 per QALY gained, based on current guidelines for health technology appraisal from the National Institute for Health and Clinical Excellence (NICE). As PCV13 and PCV15 are currently administered in a 1+1 dosing schedule in the UK, the model also considered PCV20 in a 1+1 dosing schedule. However, an analysis which considered PCV20 in a 2+1 schedule was also included to align with the standard of care dosing schedule across other European countries.

Therefore, the model considered the following comparisons:

- PCV20 2+1 vs PCV15 1+1
- PCV20 2+1 vs PCV13 1+1
- PCV20 1+1 vs PCV15 1+1
- PCV20 1+1 vs PCV13 1+1
- PCV20 2+1 vs PCV20 1+1

#### Number Needed to Vaccinate (NNV)

To further explore the public health impact of infant PCVs, the NNV for PCV20 and PCV15 compared to PCV13 was estimated using the clinical outcomes from the epidemiologic model. The NNV is a cost agnostic metric which represents the number of people who must be vaccinated to prevent one case of disease or other disease outcome.

The NNV outcomes were estimated using the following approach described by Dawson et al[24] where:

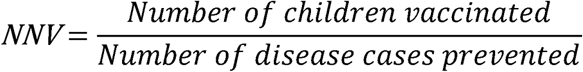

This approach intends to capture the evolving impact of pediatric pneumococcal vaccination programs over time following the accumulation of indirect effects in the unvaccinated population.

The NNV to avert a disease case (IPD, pneumonia, and OM) and pneumococcal-related death were calculated for PCV20 versus PCV13 and PCV15 versus PCV13 over a 10-year time horizon.

#### Sensitivity and scenario Analyses

Deterministic one-way and multivariate probabilistic sensitivity analyses (PSA) were employed to more thoroughly evaluate the influence of parameter uncertainty on the study outcomes. Given the two-part structure of the model, the probabilistic sensitivity analysis was conducted in distinct phases: initially, a Latin Hypercube sampling approach was applied to the parameters involved in calibrating the IPD epidemiological model, generating 1,000 samples; subsequently, a second-order Monte Carlo simulation comprising 3,000 iterations was performed, wherein the remaining parameters were varied by ±20% from their base-case estimates. Results for the PSA were presented vs PCV13.

We considered different scenarios to assess how the cost-effectiveness results may change when modifying certain parameters with higher uncertainty, which are detailed in Table 2.

**Table 2.** Scenario analyses.

| Parameter | Base-case | Scenario analysis |
| --- | --- | --- |
| PCV15 and PCV20 VE<sub>c</sub> against PCV13 serotypes | Assumed to be equivalent to PCV13 VE<sub>c</sub> | Reduced by a proportion of 21% in alignment with Choi et al[4]. |
| Time horizon | 10 years | 5 years |
| IPD presentation | Age-specific estimates of the proportion of IPD presenting as meningitis, based on data from Oligbu et al[13] | 6.5% of IPD presents as meningitis for all ages, based on data from Bertran et al[3] |

IPD: invasive pneumococcal disease; PCV: pneumococcal conjugate vaccine.

## Results

### Model calibration

Figures 2 and 3 present graphical representation of the calibration results comparing the modeled IPD incidence over time with the observed IPD incidence over time for each serotype group in ages <5 years and 65+ years (the age groups with the highest IPD incidence historically). Similar graphs for the remaining age groups can be found in the supplementary appendix, Figures S1-S3. Visual inspection and quantitative analysis suggest the model fit well to historical data. The coefficients of determination for each age group were as follows: 0.978 for ages <5 years; 0.905 for ages 5-14 years; 0.911 for ages 15-44 years; 0.893 for ages 45-64 years; and 0.820 for ages 65+ years.

**Figure 2.**
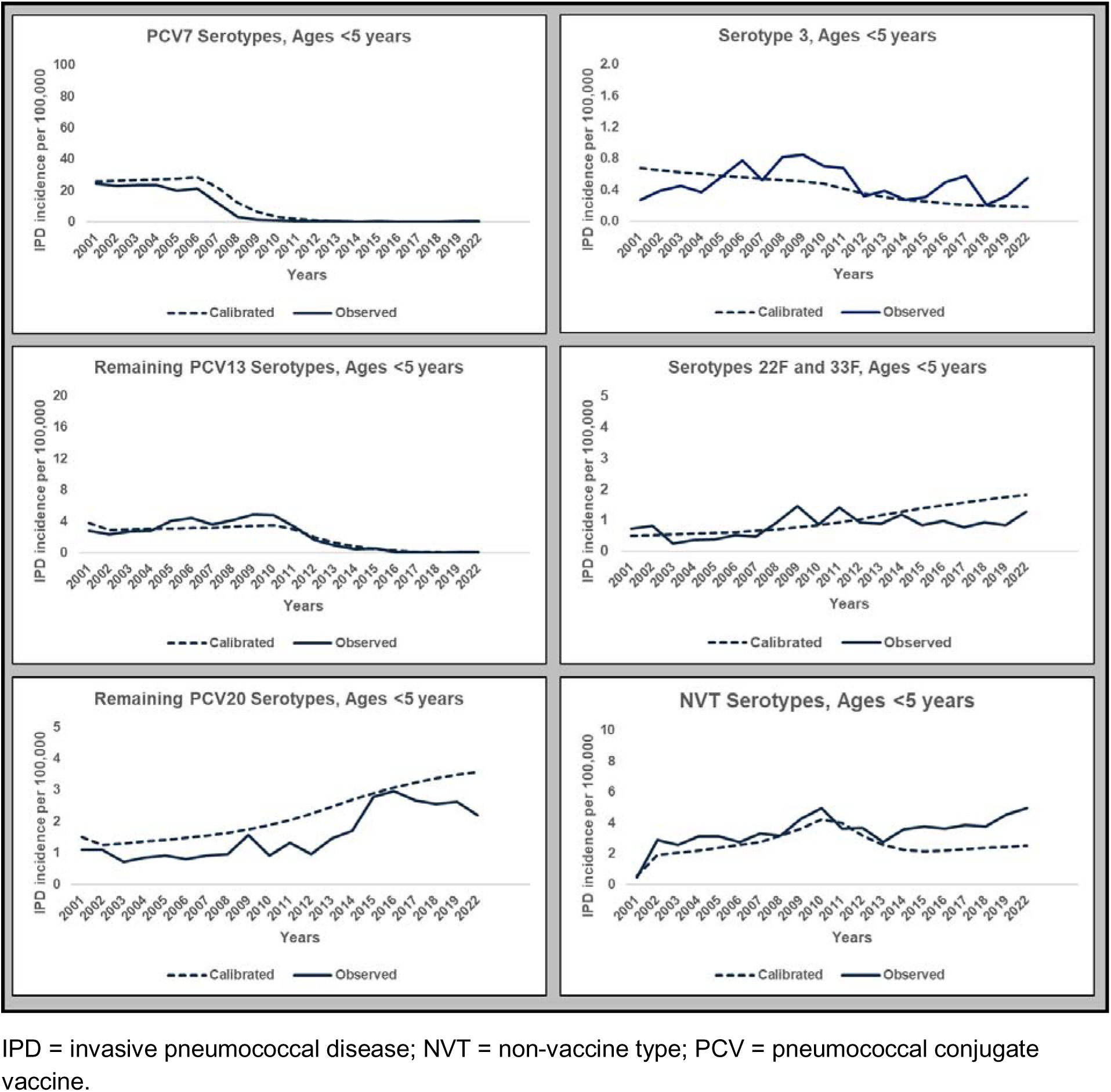
Calibrated IPD incidence results for ages <5 years.

**Figure 3.**
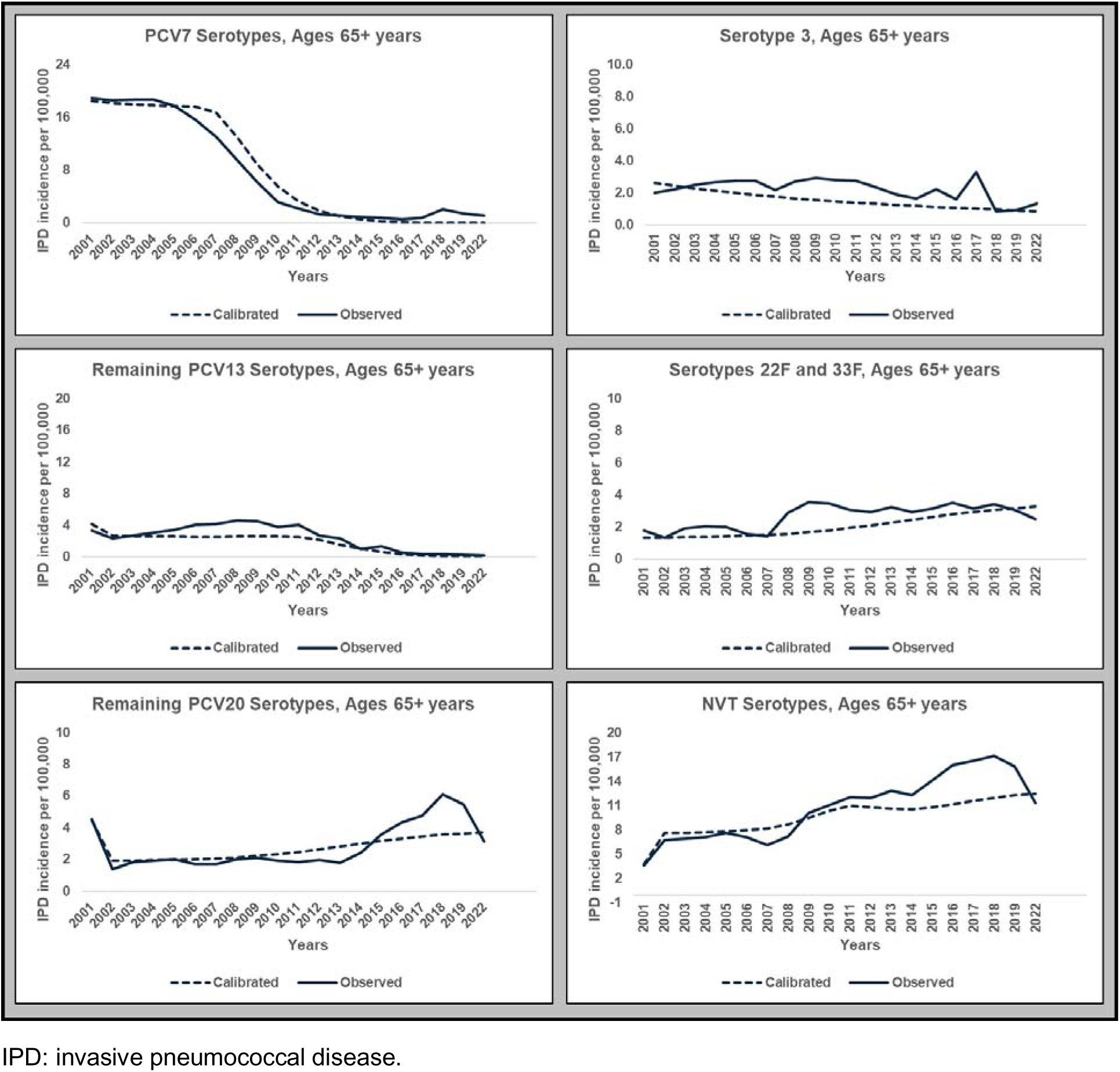
Calibrated IPD incidence results for ages 65+ years.

### Cost-effectiveness analysis

#### Base-case analysis

Over a 10-year time-horizon, PCV20 1+1 and PCV20 2+1 were estimated to be dominant (both cost-saving and more effective) strategies compared to both PCV13 1+1 and PCV15 1+1 (Table *2*, Table 3). Compared with PCV15 1+1 and PCV13 1+1, PCV20 1+1 was anticipated to avert 288,688 and 309,123 cases of disease over 10 years, respectively, at a cost saving of £1.596 billion and £1.699 billion, respectively. Similarly, PCV20 2+1 was expected to avert 494,491 and 536,570 disease cases compared with PCV15 1+1 and PCV13 1+1, respectively, with cost savings of £1.418 and £1.521 billion, respectively. When comparing the two PCV20 dosing schedules, a 2+1 PCV20 schedule was expected to avert an additional 55,130 cases of disease, with an incremental cost-effectiveness ratio of £13,656 per QALY gained.

**Table 3.** Base-case cost-effectiveness results over a 10-year time horizon.

| Parameter | PCV13 1+1 | PCV15 1+1 | PCV20 1+1 | PCV20 2+1 |
| --- | --- | --- | --- | --- |
| **Cases** |  |  |  |  |
| Bacteremia | 48,346 | 47,681 | 37,932 | 36,605 |
| Meningitis | 8,199 | 8,086 | 6,433 | 6,208 |
| Pneumonia | 1,572,758 | 1,552,323 | 1,263,635 | 1,222,764 |
| OM | 615,588 | 595,519 | 460,794 | 448,454 |
| Total | 2,265,980 | 2,223,901 | 1,784,540 | 1,729,410 |
| **Deaths** |  |  |  |  |
| IPD | 9,047 | 8,929 | 7,208 | 6,967 |
| Pneumonia | 472,749 | 466,770 | 384,656 | 372,752 |
| **Outcomes** |  |  |  |  |
| Life-years | 536,228,127 | 536,234,112 | 536,316,585 | 536,328,525 |
| QALYs | 391,909,775 | 391,916,032 | 392,005,657 | 392,018,685 |
| **Costs (£)** |  |  |  |  |
| Vaccination | 714,719,619 | 729,216,975 | 804,119,978 | 1,218,206,654 |
| IPD | 321,914,019 | 317,847,208 | 253,551,236 | 244,680,243 |
| Pneumonia | 8,660,580,846 | 8,551,849,867 | 6,975,045,529 | 6,750,448,810 |
| OM | 138,151,689 | 133,533,914 | 103,370,944 | 100,660,520 |
| Total | 9,835,366,173 | 9,732,447,964 | 8,136,087,687 | 8,313,996,228 |
| **ICER (£/QALY)** |  |  |  |  |
| vs. PCV13 1+1 | -- | PCV15<br>dominant | PCV20<br>dominant | PCV20<br>dominant |
| vs. PCV15 1+1 | -- | -- | PCV20<br>dominant | PCV20<br>dominant |
| vs. PCV20 1+1 | -- | -- | -- | 13,656 |

IPD: invasive pneumococcal disease; OM: otitis media; PCV: pneumococcal conjugate vaccine; GBP: Great British Pound; QALY: quality-adjusted life-year

#### Scenario analyses

Table 4 presents the scenario analysis results for the comparison between PCV20 1+1 and PCV15 1+1. Table 5 presents the scenario analysis results for PCV20 2+1 vs all other comparators. Each scenario analysis resulted in similar findings to the base case analysis.

**Table 4.** Scenario analysis results for PCV20 2+1 comparisons.

| Parameter | PCV20 1+1 vs PCV13 1+1 |  |  | PCV20 1+1 vs PCV15 1+1 |  |  |
| --- | --- | --- | --- | --- | --- | --- |
|  | Incr. Cost (£) | Incr. QALY | ICER (£/QALY) | Incr. Cost (£) | Incr. QALY | ICER (£/QALY) |
| Base case | -<br>1,699,278,486 | 95,882 | PCV20<br>dominant | -<br>1,596,360,277 | 89,626 | PCV20<br>dominant |
| *Scenario analyses* |  |  |  |  |  |  |
| PCV15 and PCV20 VEC against PCV13 serotypes reduced by 21% in alignment with Choi et al[4]. | -<br>1,699,223,055 | 95,880 | PCV20<br>dominant | -<br>1,596,295,762 | 89,622 | PCV20<br>dominant |
| Time horizon of 5 years | -624,201,474 | 38,064 | PCV20<br>Dominant | -603,247,630 | 36,472 | PCV20<br>dominant |
| Assuming 6.5% of IPD presents as meningitis for all ages [3] | -<br>1,699,287,990 | 95,882 | PCV20<br>dominant | -<br>1,596,375,453 | 89,625 | PCV20<br>dominant |

IPD: invasive pneumococcal disease; PCV = pneumococcal conjugate vaccine.
Note: base case considered a 10-year horizon, assumed VEC against PCV13 serotypes for PCV15 and PCV20 to be equal to that of PCV13, and assumed the distribution of IPD from Oligbu et al[13].

**Table 5.** Scenario analysis results for PCV20 2+1 comparisons.

| Parameter | PCV20 2+1 vs PCV13 1+1 |  |  | PCV20 2+1 vs PCV15 1+1 |  |  | PCV20 2+1 vs PCV20 1+1 |  |  |
| --- | --- | --- | --- | --- | --- | --- | --- | --- | --- |
|  | Incr. Cost (£) | Incr. QALY | ICER (£/QALY) | Incr. Cost (£) | Incr. QALY | ICER (£/QALY) | Incr. Cost (£) | Incr. QALY | ICER (£/QALY) |
| Base case | -<br>1,521,369,946 | 108,910 | PCV20<br>dominant | -<br>1,418,451,737 | 102,654 | PCV20<br>dominant | 177,908,541 | 13,028 | 13,656 |
| *Scenario analyses* |  |  |  |  |  |  |  |  |  |
| PCV15 and PCV20<br>VE<sub>c</sub> against PCV13<br>serotypes reduced by<br>21% in alignment with<br>Choi et al[4]. | -<br>1,521,302,962 | 108,907 | PCV20<br>dominant | -<br>1,418,375,669 | 102,650 | PCV20<br>dominant | 177,920,094 | 13,027 | 13,657 |
| Time horizon of 5<br>years | -508,431,334 | 44,365 | PCV20<br>dominant | -487,484,108 | 42,774 | PCV20<br>dominant | 115,763,522 | 6,302 | 18,370 |
| Assuming 6.5% of IPD<br>presents as meningitis<br>for all ages [3] | -<br>1,521,385,058 | 108,910 | PCV20<br>dominant | -<br>1,418,472,520 | 102,653 | PCV20<br>dominant | 177,902,933 | 13,028 | 13,655 |

IPD: invasive pneumococcal disease; PCV = pneumococcal conjugate vaccine.
Note: base case considered a 10-year horizon, assumed VE<sub>c</sub> against PCV13 serotypes for PCV15 and PCV20 to be equal to that of PCV13, and assumed the distribution of IPD from Oligbu et al., 2019.

#### One-way sensitivity analyses

Figure 4 and Figure 5 present the one-way analysis results for the economic model parameters. For both the comparison of PCV20 1+1 vs PCV13 1+1 and PCV20 2+1 vs PCV13 1+1, the results were most sensitive to the variation in inputs for pneumonia in the elderly (direct costs, case fatality rate, utility decrement) and the baseline utility for the elderly. However, in all economic parameter variation, PCV20 remained dominant.

**Figure 4.**
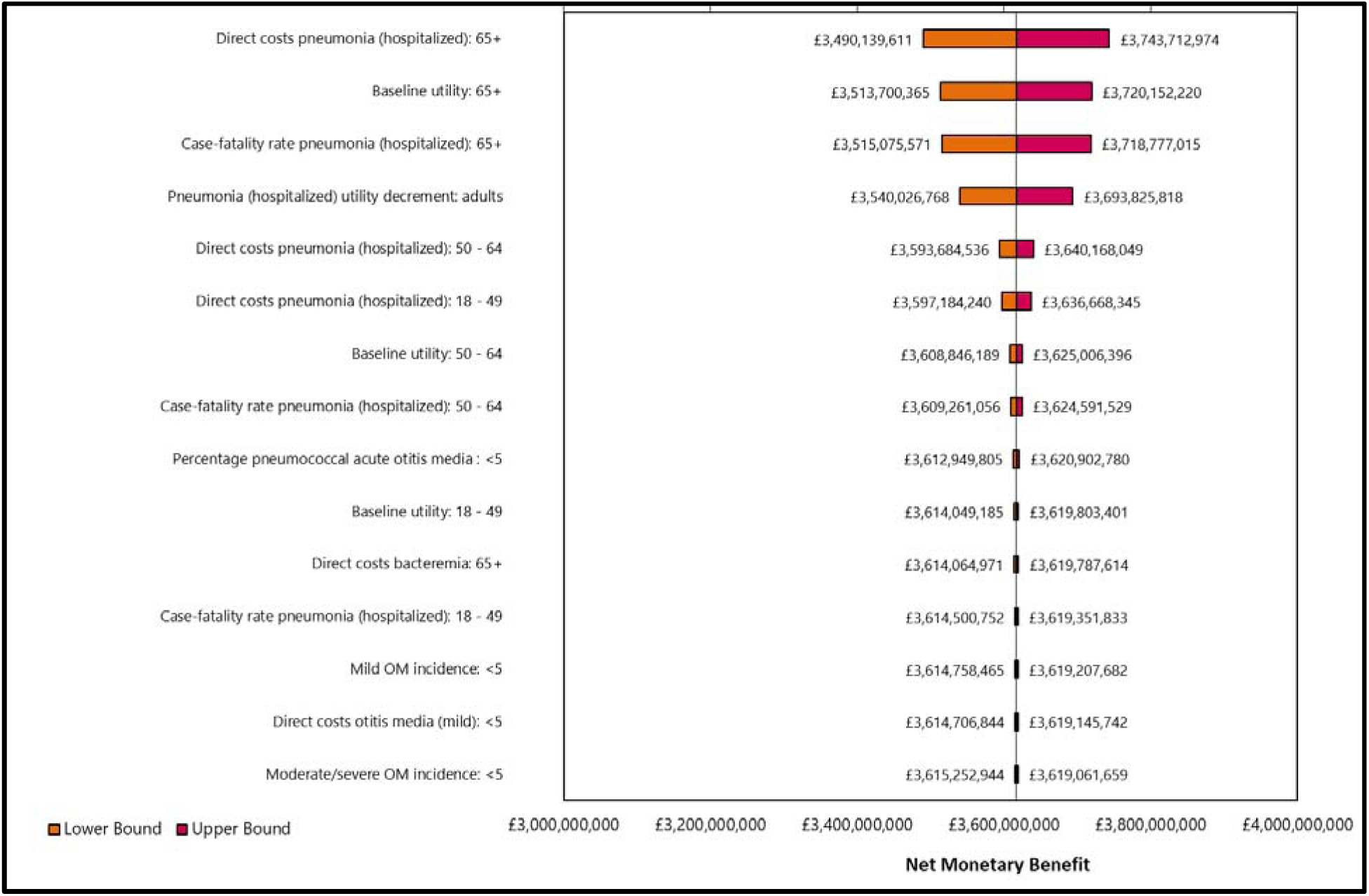
One-way sensitivity analysis results comparing PCV20 1+1 vs PCV13 1+1.

**Figure 5.**
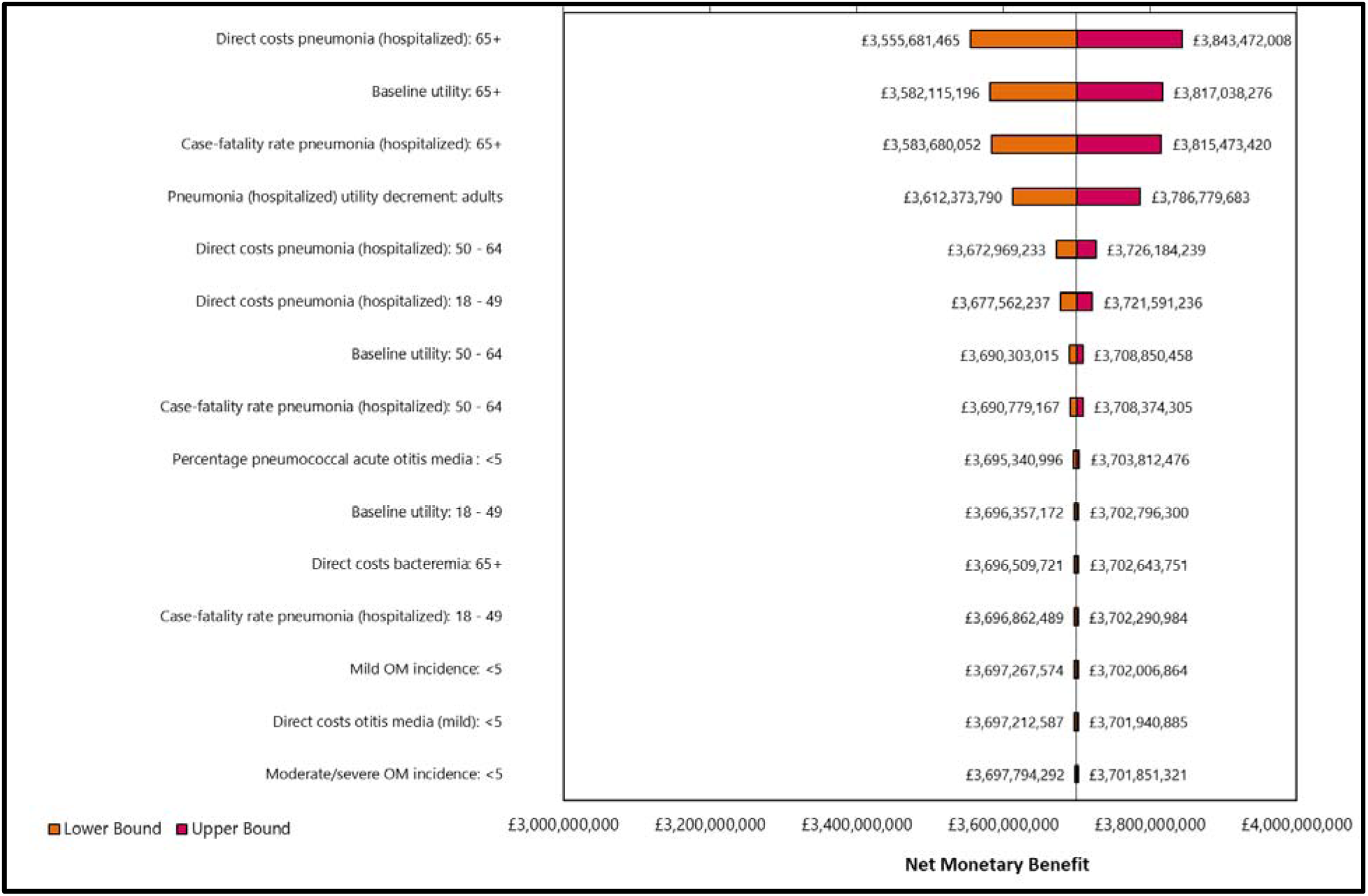
One-way sensitivity analysis results comparing PCV20 2+1 vs PCV13 1+1.

#### Probabilistic sensitivity analyses

Figure 5 and 6 present the PSA results for the economic model parameters in the form of an x/y scatter plot. Tables 5-7 present the tabular results of the Latin hypercube sampling for the calibrated epidemiologic model parameters.

**Figure 6.**
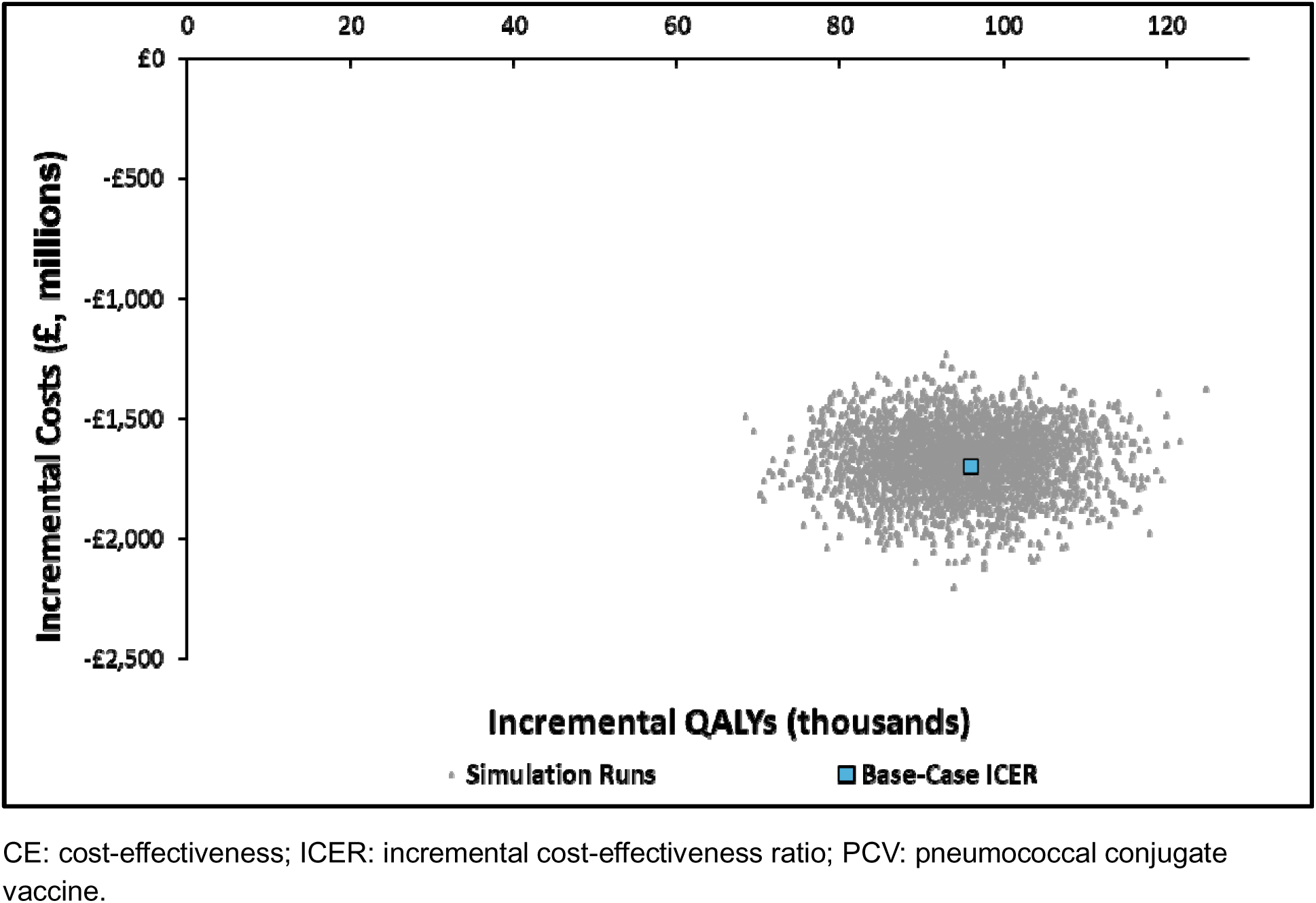
Probabilistic sensitivity analysis results comparing PCV20 1+1 vs PCV13 1+1.

**Figure 7.**
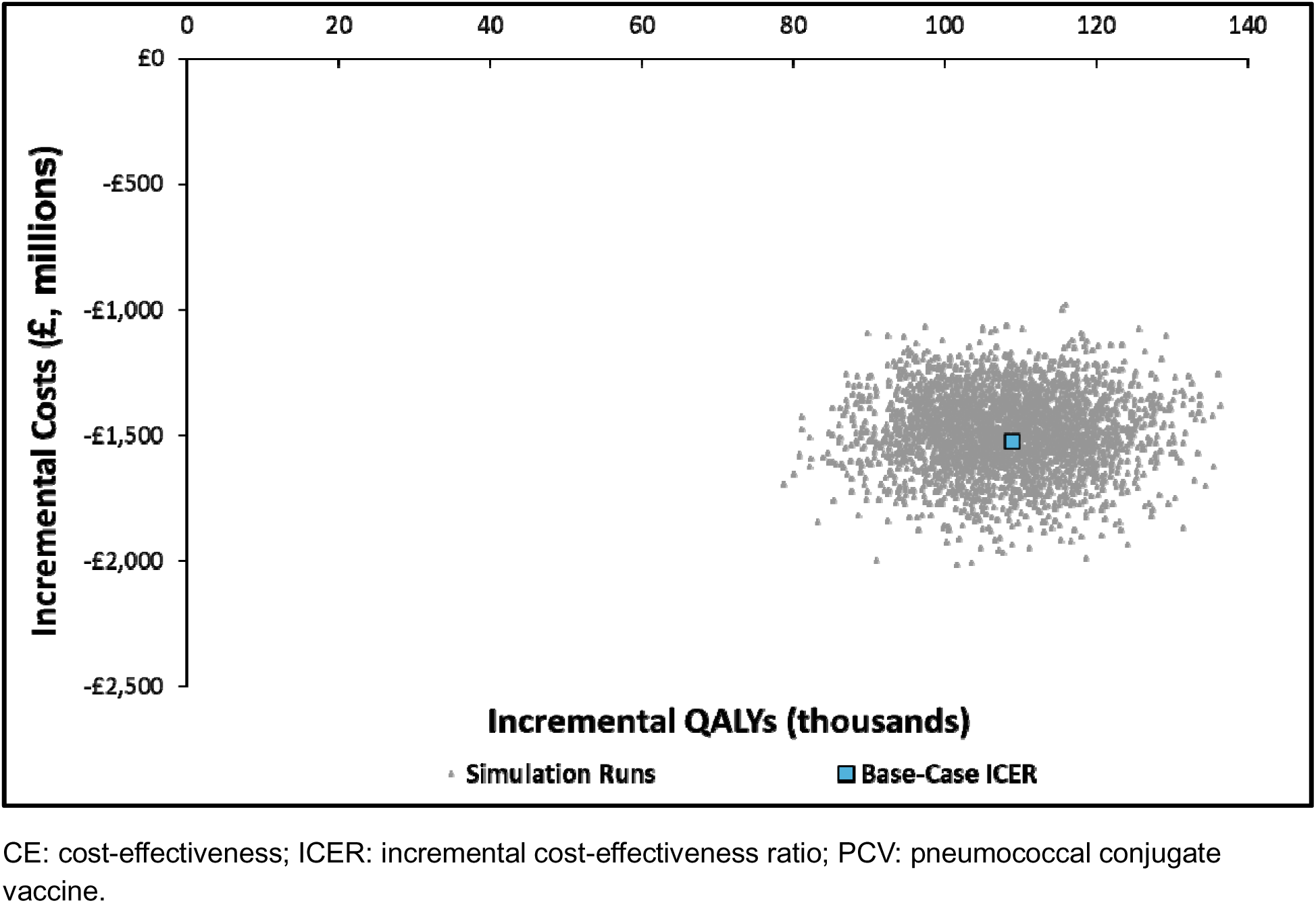
Probabilistic sensitivity analysis results comparing PCV20 2+1 vs PCV13 1+1.

**Table 6.** NNV to prevent a case of pneumococcal disease and a pneumococcal related death.

|  | **PCV15 1+1**<br>**vs PCV13**<br>**1+1** | **PCV20 1+1**<br>**vs PCV13**<br>**1+1** | **PCV20 2+1 vs**<br>**PCV13 1+1** |
| --- | --- | --- | --- |
| IPD case | 9,352 | 597 | 529 |
| Pneumonia case | 356 | 24 | 21 |
| OM case | 348 | 45 | 42 |
| Pneumococcal-related death | 1192 | 81 | 71 |

IPD: invasive pneumococcal disease; NNV: number needed to vaccinate; OM: otitis media; PCV: pneumococcal conjugate vaccine.

In the PSA of the economic parameters, PCV20 1+1 remained dominant in 100% of simulations compared with PCV13 1+1 (Figure 5). Similarly, PCV20 2+1 was dominant in all simulations vs PCV13 1+1.

Results of the Latin hypercube sampling were generally consistent with the base-case results (Table S5). When comparing PCV20 1+1 vs PCV15 1+1 or PCV13 1+1 (Table S6), PCV20 1+1 remained dominant (cost-saving and more effective) versus either lower-valent vaccine at the 95% confidence level. Similarly, PCV20 2+1 remained dominant when compared with PCV15 1+1 or PCV13 1+1 (Table S7). When comparing PCV20 2+1 to PCV15 1+1, the 95% confidence interval for the ICER ranged from being dominant to an ICER of £26,292/QALY (PCV20 2+1 generally being the cost-effective strategy).

### Number needed to vaccinate (NNV)

Table 8 presents the NNV to prevent one case of pneumococcal disease or one pneumococcal-related death for PCV15 1+1, PCV20 2+1 and PCV20 1+1 compared with PCV13 1+1.

In a 1+1 schedule, the NNV to prevent a case of IPD, pneumonia, and OM with PCV20 were 597, 24, and 45. The NNV for PCV20 2+1 were 11%, 12%, and 7% lower than those for PCV20 in a 1+1 schedule. However, regardless of the PCV20 schedule (1+1 or 2+1), the NNV to prevent a single case of IPD compared to PCV13 (1+1) was about 94% lower than with PCV15 (597 and 529 vs 9,352). Likewise, the NNVs for preventing one case of pneumonia and OM with PCV20 were roughly 93% lower (24 and 21 vs 356) and 87% lower (45 and 42 vs 348), respectively, than those for PCV15.

Similarly, the NNV to prevent one pneumococcal-related death was also nearly 15-17 times greater with PCV15 1+1 compared to PCV20 1+1 and 2+1 relative to PCV13 (1,192 vs 81 and 71). Additionally, 10 fewer children need to be vaccinated with PCV20 in a 2+1 schedule (71) compared to PCV20 in a 1+1 schedule (81) to prevent one pneumococcal-related death.

## Discussion

This is the first study to assess the cost-effectiveness and estimate the NNV for PCV20 in both the 1+1 and 2+1 dosing schedules relative to PCV13 and PCV15 using post-COVID19 pandemic epidemiologic data. Over 10 years, PCV20 1+1 and 2+1 were estimated to be dominant strategies compared to PCV13, preventing 309,000 and 536,000 additional pneumococcal disease cases and saving an additional £1.7 and £1.5 billion, respectively. Similarly, PCV20 1+1 and 2+1 were also estimated to dominate PCV15, preventing 288,000 and 494,000 more cases and saving an additional £1.6 and £1.4 billion, respectively. Across all clinical outcomes evaluated, the NNV with PCV20 1+1 was approximately 87% lower than that for PCV15 relative to PCV13, while the NNV with PCV20 2+1 was 94% lower than those estimated for PCV15 compared to PCV13, indicating greater population level efficiency of disease prevention. Collectively, these findings indicate that PCV20 under both a 1+1 and 2+1 schedule could offer meaningful clinical benefits that are accompanied by favorable economic returns relative to lower-valent alternatives.

These analyses add to the growing evidence that PCV20 is a dominant intervention compared to PCV13 and PCV15 across multiple dosing schedules and socioeconomic settings [25]. Additionally, the conclusions of this analysis are aligned with our previous estimations of PCV15 and PCV20’s impact and cost-effectiveness in the UK, which leveraged epidemiologic data through 2018 for model calibration [5]. However, the current analysis projected a substantially higher number of disease cases under all vaccine strategies, consistent with increases in disease incidence, as well as higher direct medical costs, reflecting higher costs per disease state. Though our analysis did not predict that PCV15 implementation would increase disease incidence as reported in Choi et al., the incremental benefit of PCV15 was estimated to be marginal compared to PCV13, specifically for IPD [4]. Taken together, these data support the plausibility of high public health impact and economic returns for pediatric PCV20 implementation in the UK.

Importantly, there are few dynamic models which assess the cost-effectiveness of PCV20 relatively to PCV13 and PCV15 as most of the published literature of higher-valent PCV economic evaluations employ static Markov model frameworks. While there are limited cost-effectiveness analyses in European settings employing a dynamic model framework, one dynamic model cost-effectiveness analysis conducted in the United States (US) comparing PCV20 with PCV15 and PCV13 under a 3+1 dosing schedule also used a similar model structure [26]. Despite differences in population and vaccination schedule between the UK and the US, the US-based analysis similarly demonstrated the dominance of PCV20 relative to PCV13 and PCV15. A modeling study in Israel, South Africa, and Malawi found PCV20 to be consistently substantially more effective in preventing IPD cases in ages <5 years, with PCV15 providing minimal incremental benefit over PCV13 [27]. The consistent directional findings across modeling approaches and geographies underscore the greater public health potential impact that PCV20 could have.

Furthermore, our NNV estimates for PCV20 and PCV15 are consistent with those presented in a recent analysis which estimated PCV20 and PCV15 NNVs in 21 countries, in which PCV20 consistently had lower NNVs than PCV15 relative to PCV13. This similarity is notable given differences in epidemiology, baseline disease burden, and immunisation programmes across countries, suggesting that the benefit of PCV20’s broader coverage is not highly sensitive to country-specific assumptions [24]. Additionally, the NNVs presented in this analysis are representative only of the cases prevented due to the additional serotypes covered by PCV15 and PCV20 because they were calculated relative to PCV13, which suggests the overall benefit of higher-valent vaccines may be greater than estimated. Taken together, this evidence further supports PCV20 as an economically efficient option for maximising public health benefits.

Although the UK has used a 1+1 dosing schedule in the paediatric NIP since 2020, the analysis also considered PCV20 in a 2+1 schedule and directly compared the two PCV20 dosing strategies. In the head-to-head comparison, PCV20 2+1 averted an additional 55,130 disease cases relative to PCV20 1+1 at an ICER of £13,656 per QALY gained, which falls below conventional JCVI/ NICE willingness-to-pay thresholds, suggesting the additional priming dose represents a cost-effective investment. Despite the cost of the additional dose, PCV20 2+1 was also estimated to be a cost-saving strategy versus PCV13 and PCV15 1+1. This is likely due to increased effectiveness against carriage in the first year of life, resulting in both improved direct protection for infants and indirect protection for adults [8]. Evidence from both PCV7 and PCV13 indicated that administering additional priming doses leads to increased effectiveness during the first year of life [8,28], so the removal of a priming series dose may have unintended negative consequences. While more robust data on the effectiveness of PCV20 in both schedules are needed, the model results suggest that PCV20 2+1 could provide enhanced public health benefits relative to PCV20 1+1. Decision-makers should weigh the incremental clinical gains of a 2+1 schedule against the programmatic simplicity of a 1+1 schedule.

Our analysis likely underestimates the full value of PCV20 by not capturing several downstream benefits. Pneumococcal disease, particularly OM in children, is a major driver of antibiotic prescribing in primary care, and by preventing a substantial number of disease episodes, broader-valent vaccines could reduce antibiotic consumption and associated selection pressure for resistant organisms – an important consideration given the UK’s national commitment to tackling antimicrobial resistance [29,30]. Additionally, the model captured only acute disease episodes and short-term QALY decrements but did not account for the chronic burden of pneumococcal meningitis-related sequelae such as hearing loss, cognitive impairment, and developmental delay, which impose substantial lifetime QALY losses and ongoing healthcare costs. Incorporating both AMR-related benefits and long-term sequelae into future assessments would provide a more complete picture of PCV20’s value and could further strengthen the case for broader serotype coverage.

Similar to all epidemiologic modelling efforts, our model, upon which the three analyses are based, is subject to limitations, mainly the absence of VE data against carriage and disease outcomes for PCV15 and PCV20. Consequently, our base-case analyses relied on VE estimates derived from PCV13 under the assumption that effectiveness for PCV15 and PCV20 would be comparable for the shared serotypes. This approach is consistent with prior pneumococcal DTMs and economic models evaluating newly introduced PCVs [5,26]. However, assumptions regarding protection against carriage of vaccine-type serotypes are a critical driver of indirect effects and overall population impact in our model. Therefore, we explored a scenario in which PCV15 and PCV20 were assumed to have reduced effectiveness against carriage of PCV13 serotypes, consistent with the assumptions used by Choi et al[4]. In this scenario, a greater number of cases caused by PCV13 serotypes was observed in both the PCV15 and PCV20 arms, ultimately attenuating the impact of both direct and indirect effects. However, this did not change the overall conclusions of the analysis, with results remaining closely aligned with the base case.

An additional limitation is that information regarding emerging serotypes following PCV15 and PCV20 implementation are currently unknown. Considering this uncertainty, the model assumed that future replacement patterns will resemble historical trends observed following PCV7 and PCV13 introduction. Following PCV7 introduction, certain serotypes were common among multiple geographic settings. However, the serotype landscape following introduction of higher-valent vaccines such as PCV13 resulted in a divergence of the serotype landscape such that there are no specific dominating serotypes in all countries [31]. As such, the ecological behaviour of emerging serotypes may differ following PCV15 and PCV20 implementation, potentially affecting predictions of disease burden and cost11effectiveness. An inherent limitation of pneumococcal dynamic transmission models is limited data on contact matrices or duration of carriage. This analysis leveraged a relatively old contact matrix which may not be reflective of current transmission dynamics, particularly in a post-COVID19 pandemic world. However, as this input is integral for completing the model calibration, we were unable to determine the impact that this limitation could have on the results. The model also conservatively excluded outpatient pneumonia and assumed a relatively low proportion of all-cause hospitalised pneumonia was due to pneumococcus, both of which may result in an underestimation of benefit across all modelled vaccines.

Finally, our analysis estimated population-level health and economic impacts but did not examine how the benefits of PCV20 may be distributed across socioeconomic, ethnic, or geographic subgroups within the UK. Evidence suggests that disadvantaged communities bear a disproportionate burden of pneumococcal disease due to factors such as crowded housing, higher rates of comorbidities, and inequitable access to healthcare [32]. Broader serotype coverage through PCV20 may therefore confer greater absolute benefits in these populations, potentially narrowing health disparities. As health equity considerations are increasingly central to JCVI deliberations, future work should assess the distributional impact of higher-valent PCVs across population subgroups. Despite these limitations, the study provides a comprehensive assessment of PCV20’s expected value in the UK if adopted into the paediatric NIP.

## Conclusion

The cost-effectiveness and NNV results suggest that incorporating paediatric PCV20 in the current UK pneumococcal NIP could deliver substantial public health gains and enhanced quality of life for all ages in the UK while saving costs. Importantly, these benefits are maintained under conservative assumptions and alternative scenarios, underscoring the robustness of the conclusions. These findings indicate that continued reliance on PCV13 could result in public health and economic consequences which otherwise could be avoided. Timely implementation of PCV20 in the UK paediatric NIP is warranted to optimise disease prevention, improve health system efficiency, and strengthen population11level protection against pneumococcal disease.

## Author Contributions

Methodology: Sophie Waren, Jack Said, Joe Trim, Euan Dawson, Michele Wilson, Benjamin M. Althouse, and Mark Rozenbaum; Formal analysis: Euan Dawson and Michele Wilson; Writing – Review & Editing: Sophie Waren, Jack Said, Joe Trim, Euan Dawson, Michele Wilson, Benjamin M. Althouse, and Mark Rozenbaum.

## Data Availability Statement

All data generated or analyzed during this study are included in this published article/as supplementary information files.

