## Supplement for "Projected health and economic impact of PCV20 vaccination in UK children: a dynamic transmission model"

### Supplementary Materials

#### Direct medical cost calculations per manifestation

Average unit costs for bacteremia, meningitis, hospitalized pneumonia, and otitis media were obtained from the 2022/2023 National Schedule of NHS Costs to align with the date of the latest model-calibrated IPD incidence data from Bertran et al. 2024.

International Classification of Diseases, Tenth Revision (ICD-10) codes for each manifestation were mapped to relevant Healthcare Resource Group (HRG) codes and, for each HRG code, activity-weighted national average unit costs per finished consultant episode (FCE) were derived. Where multiple HRG codes were relevant for a single manifestation, a weighted average of the individual HRG costs was taken to generate a single value. Unit costs for bacteremia, meningitis, and pneumonia were reported separately for adults and pediatrics, based on their unique HRG codes. The costs for otitis media were reported for pediatrics only and broken down into moderate/severe cases (admitted attendance) and mild cases (outpatient attendance).

ICD-10 codes for each manifestation were mapped to HRG codes as follows:

- **Bacteremia:** ICD-10 codes A40.3, A40.8, A40.9, A41.9, A49.1, A49.9, and I33.0 were mapped to HRG codes WJ06, WJ03, WJ02, and EB02 for adult patients and PW16, PW17, and PE23 for pediatric patients.
- **Meningitis:** ICD-10 codes G00.1, G00.8, G00.9, G04.2, G04.8, G04.9, and G05.0 were mapped to HRG code AA22 for adult patients and PW16 for pediatric patients.
- **Pneumonia:** ICD-10 codes J13, J15.9, and J18 were mapped to HRG codes DZ11 and DZ23 for adult patients and PD14 for pediatric patients.
- **Otitis media (moderate/severe cases):** ICD-10 codes H65.0, H65.1, H65.2, H65.3, H65.4, H65.9, H66.0, H66.1, H66.2, H66.3, H66.4, H66.9, H67.0, H67.1, H67.8 were mapped to PC63 for pediatric patients.
- **Otits media (mild cases):** The unit cost for a pediatric outpatient attendance was derived from HRG code CA54B for Minor Ear Procedures, 18 years and under.

For each manifestation, the average cost per hospital admission was then calculated by multiplying the average unit cost per FCE by the average number of FCEs per admission, derived from NHS Digital’s Hospital Admitted Patient Care Activity 2022-23 (NHS, 2023). First, we derived the average number of FCEs per admission for each manifestation, based on their relevant ICD-10 codes. Secondly, the average number of FCEs per admission was multiplied by the unit cost per FCE to generate the average cost per admission. For children, the number of FCE’s per admission was assumed to equal 1 given the estimated median length of stay in hospital of 1 day in the British Thoracic Society 2016/2017 audit (Legg et al., 2018). The final cost inputs used in the model are presented in Tables S1 and S2.

**Table S1. Medical costs per manifestation for adult patients.**

|  | **Bacteremia** | **Meningitis** | **Pneumonia** |
| --- | --- | --- | --- |
| **ICD-10 codes** | A40.3, A40.8, A40.9, A41.9, A49.1, A49.9, I33.0 | G00.1, G00.8, G00.9, G04.2, G04.8, G04.9, G05.0 | J13, J15.9, J18 |
| **HRG codes** | WJ06, WJ03, WJ02, EB02 | AA22 | DZ11, DZ23 |
| **Cost per FCE** | £2,990 | £3,089 | £2,633 |
| **No. FCEs per admission** | 2.1 | 2.1 | 2.2 |
| **Cost per admission** | £6,311 | £6,476 | £5,740 |

**Table S2. Medical costs per manifestation for pediatric patients.**

|  | **Bacteremia** | **Meningitis** | **Pneumonia** | **Otitis media** |
| --- | --- | --- | --- | --- |
| **ICD-10 codes** | A40.3, A40.8, A40.9, A41.9, A49.1, A49.9, I33.0 | G00.1, G00.8, G00.9, G04.2, G04.8, G04.9, G05.0 | J13, J15.9, J18 | H65, H66, H67 |
| **HRG codes** | PW16, PW17, PE23 | PW16 | PD14 | PC63 |
| **Cost per FCE** | £2,649 | £3,705 | £2,346 | £1,034 |
| **No. FCEs per admission*** | 1.0 | 1.0 | 1.0 | 1.0 |
| **Cost per admission** | £2,649 | £3,705 | £2,346 | £1,034 |

**assumption based on Wilson et al. 2023 which assumed that, for children, the number of FCE’s per admission was equal to 1.*

**Table S3. Calibrated Vaccine Effectiveness Against IPD for Pediatric Vaccination**

| **Parameter** | **Dose 1** | **Dose 2** | **Booster** |
| --- | --- | --- | --- |
| **PCV13 (1+1)** |  |  |  |
| PCV7 serotypes | 58.9% | 96.9% | 96.5% |
| ST3 | 21.6% | 67.4% | 68.4% |
| 19A | 69.6% | 62.6% | 77.4% |
| ST 1, 5, 6A, 7F | 87.0% | 95.8% | 98.0% |
| **PCV15 (1+1)** |  |  |  |
| PCV7 serotypes | 58.9% | 96.9% | 96.5% |
| ST3 | 21.6% | 67.4% | 68.4% |
| 19A | 69.6% | 62.6% | 77.4% |
| ST 1, 5, 6A, 7F | 87.0% | 95.8% | 98.0% |
| ST 22F and 33F | 69.6% | 62.6% | 77.4% |
| **PCV20 (1+1 and 2+1)** |  |  |  |
| PCV7 serotypes | 58.9% | 96.9% | 96.5% |
| ST3 | 21.6% | 67.4% | 68.4% |
| 19A | 69.6% | 62.6% | 77.4% |
| ST 1, 5, 6A, 7F | 87.0% | 95.8% | 98.0% |
| ST 22F and 33F | 69.6% | 62.6% | 77.4% |
| ST8 | 69.6% | 62.6% | 77.4% |
| 10A, 11A, 12F, 15B | 69.6% | 62.6% | 77.4% |

IPD = invasive pneumococcal disease; PCV = pneumococcal conjugate vaccine; ST = serotype.

**Table S4. Calibrated Vaccine Effectiveness against Carriage for Pediatric Vaccination**

| **Parameter** | **Dose 1** | **Dose 2** | **Booster** |
| --- | --- | --- | --- |
| **PCV13 (1+1)** |  |  |  |
| PCV7 serotypes | 35.2% | 72.2% | 86.3% |
| ST3 | 0.8% | 5.3% | 6.8% |
| 19A | 22.1% | 34.5% | 66.0% |
| ST 1, 5, 6A, 7F | 2.8% | 21.5% | 21.5% |
| **PCV15 (1+1)** |  |  |  |
| PCV7 serotypes | 35.2% | 72.2% | 86.3% |
| ST3 | 0.8% | 5.3% | 6.8% |
| 19A | 22.1% | 34.5% | 66.0% |
| ST 1, 5, 6A, 7F | 2.8% | 21.5% | 21.5% |
| ST 22F and 33F | 22.1% | 34.5% | 48.2% |
| **PCV20 (1+1)** |  |  |  |
| PCV7 serotypes | 35.2% | 72.2% | 86.3% |
| ST3 | 0.8% | 5.3% | 6.8% |
| 19A | 22.1% | 34.5% | 66.0% |
| ST 1, 5, 6A, 7F | 2.8% | 21.5% | 21.5% |
| ST 22F and 33F | 22.1% | 34.5% | 48.2% |
| ST8 | 22.1% | 34.5% | 48.2% |
| 10A, 11A, 12F, 15B | 22.1% | 34.5% | 48.2% |
| **PCV20 (2+1)** |  |  |  |
| PCV7 serotypes | 35.2% | 72.2% | 86.3% |
| ST3 | 0.8% | 5.3% | 6.8% |
| 19A | 22.1% | 34.5% | 66.0% |
| ST 1, 5, 6A, 7F | 2.8% | 21.5% | 21.5% |
| ST 22F and 33F | 22.1% | 34.5% | 66.0% |
| ST8 | 22.1% | 34.5% | 66.0% |
| 10A, 11A, 12F, 15B | 22.1% | 34.5% | 66.0% |

PCV = pneumococcal conjugate vaccine; ST = serotype.

Table S5. Treatment-specific results of Latin hypercube sampling sensitivity analysis of calibrated parameters

| **Parameter** | **PCV13** | **PCV15** | **PCV20** | **PCV20 2+1** |
| --- | --- | --- | --- | --- |
| Costs (£; thousands) |  |  |  |  |
| Vaccine costs | 714,720  (714,720; 714,720) | 729,217  (729,217; 729,217) | 804,120  (804,120; 804,120) | 1,218,207  (1,218,207; 1,218,207) |
| IPD costs | 334,597  (259,967; 409,227) | 329,742  (255,285; 404,200) | 261,566  (198,840; 324,292) | 251,803  (191,739; 311,867) |
| Pneumonia costs | 8,990,340  (7,107,905; 10,872,775) | 8,870,845  (6,991,668; 10,750,023) | 7,208,253  (5,581,145; 8,835,362) | 6,964,790  (5,398,242; 8,531,337) |
| OM costs | 146,301  (95,388; 197,215) | 134,139  (84,330; 183,948) | 94,935  (63,880; 125,989) | 89,191  (60,657; 117,726) |
| Total costs | 10,185,958  (8,185,364; 12,186,552) | 10,071,434  (8,071,330; 12,071,538) | 8,368,874  (6,653,096; 10,084,653) | 8,523,991  (6,873,923; 10,174,059) |
| Outcomes |  |  |  |  |
| Life-years (thousands) | 536,210  (536,109; 536,312) | 536,217  (536,115; 536,318) | 536,303  (536,214; 536,393) | 536,316  (536,230; 536,402) |
| QALY (thousands) | 391,890  (391,779; 392,001) | 391,897  (391,786; 392,008) | 391,991  (391,894; 392,089) | 392,006  (391,912; 392,099) |
| Bacteremia cases | 54,977  (42,404; 67,549) | 54,015  (41,496; 66,533) | 42,602  (32,202; 53,001) | 40,964  (31,028; 50,900) |
| Meningitis cases | 3,822  (2,948; 4,696) | 3,755  (2,885; 4,625) | 2,962  (2,239; 3,685) | 2,848  (2,157; 3,539) |
| OM cases | 22,390  (14,170; 30,609) | 20,184  (12,160; 28,209) | 14,056  (9,324; 18,787) | 13,152  (8,833; 17,471) |
| Pneumonia cases | 1,633,125  (1,287,930; 1,978,321) | 1,609,564  (1,265,260; 1,953,868) | 1,303,961  (1,007,789; 1,600,133) | 1,259,225  (974,422; 1,544,027) |
| Disease cases | 2,365,903  (1,790,770; 2,941,036) | 2,286,823  (1,719,574; 2,854,072) | 1,788,982  (1,350,374; 2,227,590) | 1,716,147  (1,300,947; 2,131,347) |
| IPD deaths | 9,398  (7,372; 11,423) | 9,257  (7,237; 11,278) | 7,432  (5,713; 9,151) | 7,167  (5,518; 8,816) |
| Pneumonia deaths | 490,547  (389,415; 591,678) | 484,225  (383,260; 585,191) | 397,849  (309,201; 486,496) | 385,029  (299,412; 470,646) |

Results of 1,000 iteration Latin hypercube sampling varying parameters across a range of +/-10% from base-case values. Means (95% confidence intervals) for each outcome for each comparator are presented.

Table S6. Incremental results of Latin hypercube sampling sensitivity analysis of calibrated parameters: PCV20 1+1

| **Parameter** | **PCV20 1+1 vs PCV13** | **PCV20 1+1 vs PCV15** |
| --- | --- | --- |
| Incremental costs (£; thousands) |  |  |
| Vaccine costs | 89,400  (89,400; 89,400) | 74,903 (74,903; 74,903) |
| IPD costs | -73,030  (-94,611; -51,449) | -68,176 (-45,956; -90,396) |
| Pneumonia costs | -1,782,087  (-2,328,119; -1,236,054) | -1,662,592 (-1,102,044; -2,223,140) |
| OM costs | -51,367  (-74,904; -27,829) | -39,205 (-15,567; -62,843) |
| Total costs | -1,817,083  (-2,400,616; -1,233,551) | -1,702,560 (-1,098,344; -2,306,775) |
| Incremental outcomes |  |  |
| Life-years (thousands) | 93  (64; 123) | 87 (56; 117) |
| QALY (thousands) | 101  (70; 133) | 94  (62; 127) |
| Bacteremia cases | -12,375  (-16,079; -8,671) | -11,413  (-7,601; -15,225) |
| Meningitis cases | -860  (-1,118; -603) | -793  (-528; -1,058) |
| OM cases | -8,334  (-12,407; -4,261) | -6,129  (-2,049; -10,208) |
| Pneumonia cases | -329,165  (-429,797; -228,532) | -305,603  (-202,228; -408,978) |
| Disease cases | -576,921  (-772,303; -381,538) | -497,841  (-298,289; -697,392) |
| IPD deaths | -1,966  (-2,558; -1,374) | -1,826  (-1,217; -2,434) |
| Pneumonia deaths | -92,698  (-122,126; -63,270) | -86,377  (-56,238; -116,515) |

Results of 1,000 iteration Latin hypercube sampling varying parameters across a range of +/-10% from base-case values. Means (95% confidence intervals) for each outcome for each comparison are presented.

Table S7. Incremental results of Latin hypercube sampling sensitivity analysis of calibrated parameters: PCV20 2+1

| **Parameter** | **PCV20 2+1 vs PCV13** | **PCV20 2+1 vs PCV15** | **PCV20 2+1 vs PCV20 1+1** |
| --- | --- | --- | --- |
| Incremental costs (£; thousands) |  |  |  |
| Vaccine costs | 503,487  (503,487; 503,487) | 488,990  (488,990; 488,990) | 414,087  (414,087; 414,087) |
| IPD costs | -82,793  (-108,043; -57,544) | -77,939  (-103,728; -52,150) | -9,763  (-14,409; -5,118) |
| Pneumonia costs | -2,025,551  (-2,661,206; -1,389,895) | -1,906,056  (-2,553,647; -1,258,464) | -243,464  (-360,880; -126,047) |
| OM costs | -57,110  (-83,614; -30,606) | -44,948  (-71,579; -18,316) | -5,743  (-9,214; -2,273) |
| Total costs | -1,661,967  (-2,342,529; -981,405) | -1,547,443  (-2,245,635; -849,251) | 155,117  (30,138; 280,095) |
| Incremental outcomes |  |  |  |
| Life-years (thousands) | 106  (72; 140) | 100  (65; 135) | 13  (7; 19) |
| QALY (thousands) | 115  (79; 152) | 108  (71; 146) | 14  (7; 21) |
| Bacteremia cases | -14,012  (-18,361; -9,663) | -13,050  (-17,489; -8,611) | -1,637  (-2,427; -847) |
| Meningitis cases | -974  (-1,276; -672) | -907  (-1,216; -599) | -114  (-169; -59) |
| OM cases | -9,237  (-13,782; -4,693) | -7,032  (-11,589; -2,475) | -903  (-1,472; -335) |
| Pneumonia cases | -373,901  (-491,366; -256,436) | -350,339  (-470,048; -230,631) | -44,736  (-66,399; -23,074) |
| Disease cases | -649,756  (-877,832; -421,681) | -570,676  (-802,364; -338,988) | -72,835  (-109,835; -35,835) |
| IPD deaths | -2,231  (-2,923; -1,538) | -2,090  (-2,796; -1,384) | -265  (-392; -137) |
| Pneumonia deaths | -105,517  (-139,650; -71,385) | -99,196  (-133,899; -64,493) | -12,819  (-19,095; -6,544) |

Results of 1,000 iteration Latin hypercube sampling varying parameters across a range of +/-10% from base-case values. Means (95% confidence intervals) for each outcome for each comparison are presented.
